# Relationship between Excess Mortality Derived from Death Counts and Age-Standardised Mortality Rates: The Case of Germany During the COVID-19 Pandemic

**DOI:** 10.64898/2026.11.07.26367468

**Authors:** Martin Sauter

## Abstract

Excess mortality can be estimated from crude death counts or from age-standardised mortality rates (ASMRs). Although both approaches are widely used, their analytical relationship and the effects of methodological choices such as standard population, age-group definition, and baseline selection are not always explicitly considered. The analytical relationship between relative excess mortality based on death counts and ASMRs was derived and both approaches were applied to mortality in Germany during the COVID19 pandemic 2020–2023. Expected mortality was estimated by linear extrapolation of pre-pandemic trends. ASMR calculations were repeated using different standard populations, age-group definitions, open-ended age groups, and reference periods. Death-count- and ASMR-based P-scores were analytically equivalent when the age distribution of the standard population corresponded to that of the population under study. Empirically, both approaches yielded closely corresponding estimates for Germany. The effect of the standard population depended on the age distribution of excess mortality and was particularly pronounced in 2020, when excess mortality was concentrated at older ages. The definition of the open-ended age group had an additional effect: aggregating ages 80+ altered the pre-pandemic ASMR trend, and consequently changed the estimated excess mortality. Variation of the pre-pandemic reference period substantially affected P-scores, but death-count- and ASMR-based estimates changed in a closely corresponding manner. Relative excess mortality derived from death counts and ASMRs is closely related, and differences between the approaches can be explained by their underlying weighting and baseline assumptions. For trend-based excess mortality estimation, the standard population, age-group definition, and baseline specification are therefore integral methodological choices. Death counts provide a simple and transparent reference measure for analyses within a single population, whereas age-standardised estimates require careful consideration of these additional assumptions.

## 1 Introduction

### 1.1 Excess Mortality and its Quantification

Excess mortality is defined as the degree to which the observed number of deaths exceeds the number expected under a counterfactual baseline. It has been widely used to assess the overall mortality impact of the COVID-19 pandemic because it is based on all-cause mortality and is therefore independent of the classification and reporting of individual causes of death.

Different mortality measures can be used for the calculation of excess mortality. The most direct approach is based on crude death counts, i.e. the absolute number of deaths in a population during a specified period. Studies comparing multiple countries use death counts, as these values are publicly available from national statistic offices and the calculation is straightforward [1], [2], [3], [4].

Alternatively, mortality can be expressed as the Crude Mortality Rate (CMR), relating the number of deaths to the corresponding population size, or as an Age-Standardised Mortality Rate (ASMR), in which age-specific mortality rates are weighted according to a defined standard population. These approaches differ substantially in their data requirements and in the assumptions required for their calculation. A smaller number of authors use these measures to quantify excess mortality during the years of the COVID-19 pandemic [5], [6], [7],

Regardless of the mortality measure used, expected mortality is commonly estimated by extrapolating mortality patterns observed during a pre-pandemic reference period which raises the question how to calculate the counterfactual number of deaths. In aging populations, death counts are rising continuously and extrapolating existing trends is a straightforward approach. Linear extrapolation makes the particularly simple assumption that the direction and magnitude of the observed trend keep the same and continue beyond the reference period.

Poisson regression is used in mortality surveillance systems such as EuroMOMO for modelling annual mortality trends [8]. For the German pre-pandemic mortality data considered here, however, the gradual increase in annual deaths results in only minimal curvature of the exponential trend implied by Poisson regression. Consequently, linear and Poisson regression yield very similar baseline estimates, with differences below 0.5% for all reference periods investigated [9].

Various national statistical offices used averages or median values of previous years rather than extrapolating an existing trend [10], [11]. In the presence of a rising mortality trend, such approaches result in a lower counterfactual baseline and consequently higher estimates of excess mortality.

More complex models implementing spline-based approaches, have been used by WHO and IHME to estimate the global mortality impact of the pandemic [1], [3]. While such models can account for nonlinear patterns within the observed period, extrapolation beyond the fitted data is also influenced by the estimated curvature. Consequently, relatively small differences in curvature near the end of the reference period may result in substantially different counterfactual trajectories and these results have been discussed intensively [12]. Linear extrapolation avoids curvature and therefore provides a simple and transparent reference model.

As age standardisation is intended to remove the influence of differences in population age structure, the effect of population ageing is removed from the mortality trend. This reveals the underlying long-term decline in age-specific mortality associated with increasing life expectancy, resulting in declining ASMR trends. Analogous to increasing trends in death counts, such declining trends can be approximated and extrapolated linearly as well [7].

This raises a fundamental methodological question: under which conditions does excess mortality derived from age-standardised mortality rates provide the same result as the substantially simpler calculation based directly on death counts? Conversely, when the two approaches differ, it is important to understand whether these differences reflect the underlying mortality pattern or arise from the choice of standard population and age-group aggregation.

The present study investigates this relationship analytically and empirically. The mathematical relationship between excess mortality derived from death counts and from ASMR is first established. The effects of standard population and age-group aggregation are then examined using mortality in Germany during the COVID-19 pandemic as a case study. Particular attention is given to the contrast between years in which excess mortality was concentrated in older age groups and years in which relative excess mortality was more broadly distributed across age groups.

## 2 Methods and Analytical Framework

### 2.1 Data Sources and Computational Methods

Annual numbers of deaths in Germany, stratified by age, were obtained from the German Federal Statistical Office (Destatis) through the GENESIS database [13], [14]. Corresponding annual population data for individual ages were obtained from the same database [15]. The mortality and population data were aggregated into the age groups required for the respective analyses. The population data used in this publication represents the Census of 2011.

All calculations were performed using Python 3 [16], with NumPy [17] and SciPy [18] used for numerical calculations and linear regression. Matplotlib [19] was used for graphical presentation. The complete datasets and Python scripts used for data processing, calculation of excess mortality and age-standardised mortality rates, and generation of the figures are provided in the accompanying GitHub-Repository [20].

### 2.2 Death-Count-Based Excess Mortality

Median value for prediction comes from the linear trend from the years 2013-2019, this fit which will serve as baseline for all other analyses. An evaluation of different trends is found in section 4.3.

I will use for the linear trends formulas referring to the start value (which is the expected number of deaths in the year 2013 *D*_*0,tot*_ and a slope m_D,tot_). The analytical expression for the trend of expected deaths is:

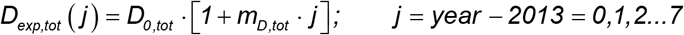

The letter “D” refers to crude death counts, the index “tot” refers to the total number of expected or observed deaths, the index will later be also used for the age groups.

The absolute excess which will be calculated from the difference between observed and expected deaths in the years between 2020 and 2023 will be named as *X*_*D,tot*_, the relative excess (later shown as percentage) will be noted in the small letter *x*_*D,tot*_ . Absolute and relative excess will both play an important role in this study.

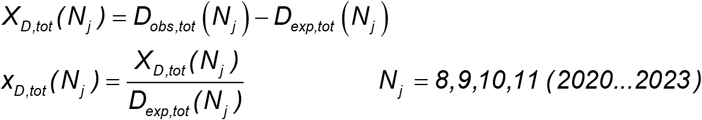

In demographic analyses the relative excess *x*_*D*_ is often referred to as “P-score”, usually this expression refers to a relative elevation in death numbers [21]. We will use the relative excess also for age-standardised mortality rates in later sections, distinguishing by another index.

### 2.3 Decomposition of Death-Count-Based Excess Mortality by Age

The total number of deaths observed in a year *j* can be expressed by the sum of deaths across *n*_*ag*_ different age groups:

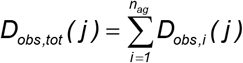

Due to the linearity of least-squares regression, a regression of the total is identical to the sum of the regressions of its subgroups. Consequently, expected and absolute excess deaths are additive across age groups:

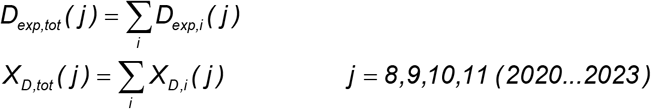

For the whole population and each age group relative and absolute excess mortality are defined as:

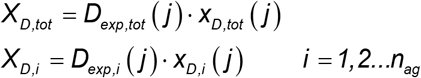

It is possible to rewrite the sum to get a relation between the relative excess numbers in each age group with the overall relative excess:

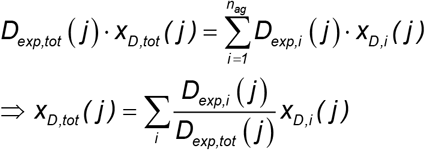

Which means that the total relative excess is the sum of the age-group specific relative excesses weighted by the proportion of expected deaths in the specific age-group to the overall expected deaths.

### 2.4 Age-Standardised Mortality Rates

Age-standardised mortality rates (ASMR) account for differences in the age structure of a population by applying age-specific mortality rates to a defined standard population.

For *n*_*ag*_ age groups, let *P*_*i*_ *( y ), i* = *1,2*,…*n*_*ag*_ denote the population size and *D*_*obs,i*_ *( j)* the observed number of deaths in age group *i* and year *j*. The corresponding age-specific crude mortality rate is

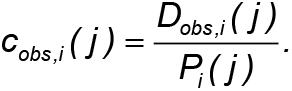

A standard population is defined by the weights *s*_*1*_,*s*_*2*_ …*s*_*n*_ with 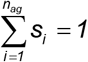. The observed age-standardised mortality rate *a*_*obs*_*(j)* is then calculated as:

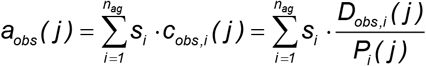

For the primary analysis, the age distribution of the German population in 2020 (DE2020) is used as the standard population, as it represents the population under study at the beginning of the investigated period. This standard population was derived from the data provided by Destatis. Alternative standard populations, including ESP2013 [22] and WHO [23], are considered subsequently to examine the effect of the choice of standard population on estimated excess mortality.

### 2.5 Analytical Relationship between Excess Mortality Derived from Death Counts and Age-Standardised Mortality Rates

Analogously, the expected age-standardised mortality rate is obtained from the expected age-specific mortality rates,

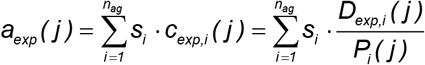

When the deaths in a year *j* and in the age group *i* exceed the expected values by the relative excess *x*_*d,i*_*(j*) the absolute difference in ASMR becomes:

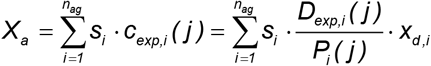

This expression is valid when using the linear fit for the death numbers for each age group *i*.

The relative excess of the ASMR value can be calculated as the relation between excess and expected value. When the fraction is expanded by the reciprocal value of the expected deaths this expression becomes similar to the P-score of deaths *x*_*d,tot*_. A second expansion by the total population number *P*_*tot*_*(j)* makes it possible to introduce a relative proportion of the population in the age groups *p*_*i*_*(j):*

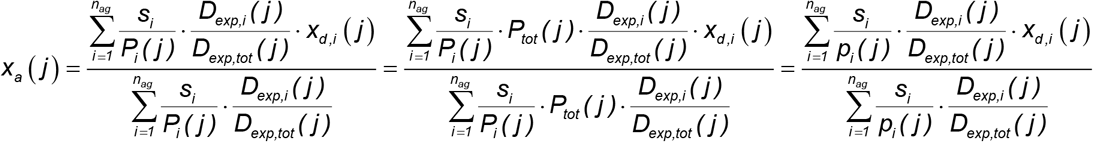

If the standard population weights s_i_ are identical to the population shares *p*_*i*_*(j)* in year *j*, the factors 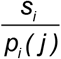 cancel. Since the age-specific expected death counts sum to the expected total death count, the denominator equals unity and

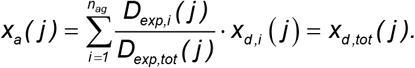

Thus, when the population under study is used as the standard population, ASMR-based and death-count-based relative excess mortality are identical in this idealised model. In real data, however, age-specific mortality fluctuates around the fitted trends, mainly due to variations in winter influenza mortality. As the expected age-specific death counts are estimated from separate regressions, these fluctuations affect the fitted trends and limit the exact correspondence between the two approaches.

## 3 Results and Discussion

### 3.1 Overall Excess Mortality in Germany derived from Death Counts and ASMR

#### 3.1.1 Overall Excess Mortality

Figure 1 shows the annual number of deaths in Germany together with the linear baseline derived from the reference period 2013–2019. During this period, annual death counts followed an approximately linear increasing trend. Relative deviations from the fitted baseline during the reference period ranged from −3.00% in 2014 to +2.06% in 2015. Among the seven reference periods ranging from 2010–2019 to 2016–2019, the 2013–2019 period yields the median baseline projection and was therefore used throughout the analysis [9].

**Figure 1.**
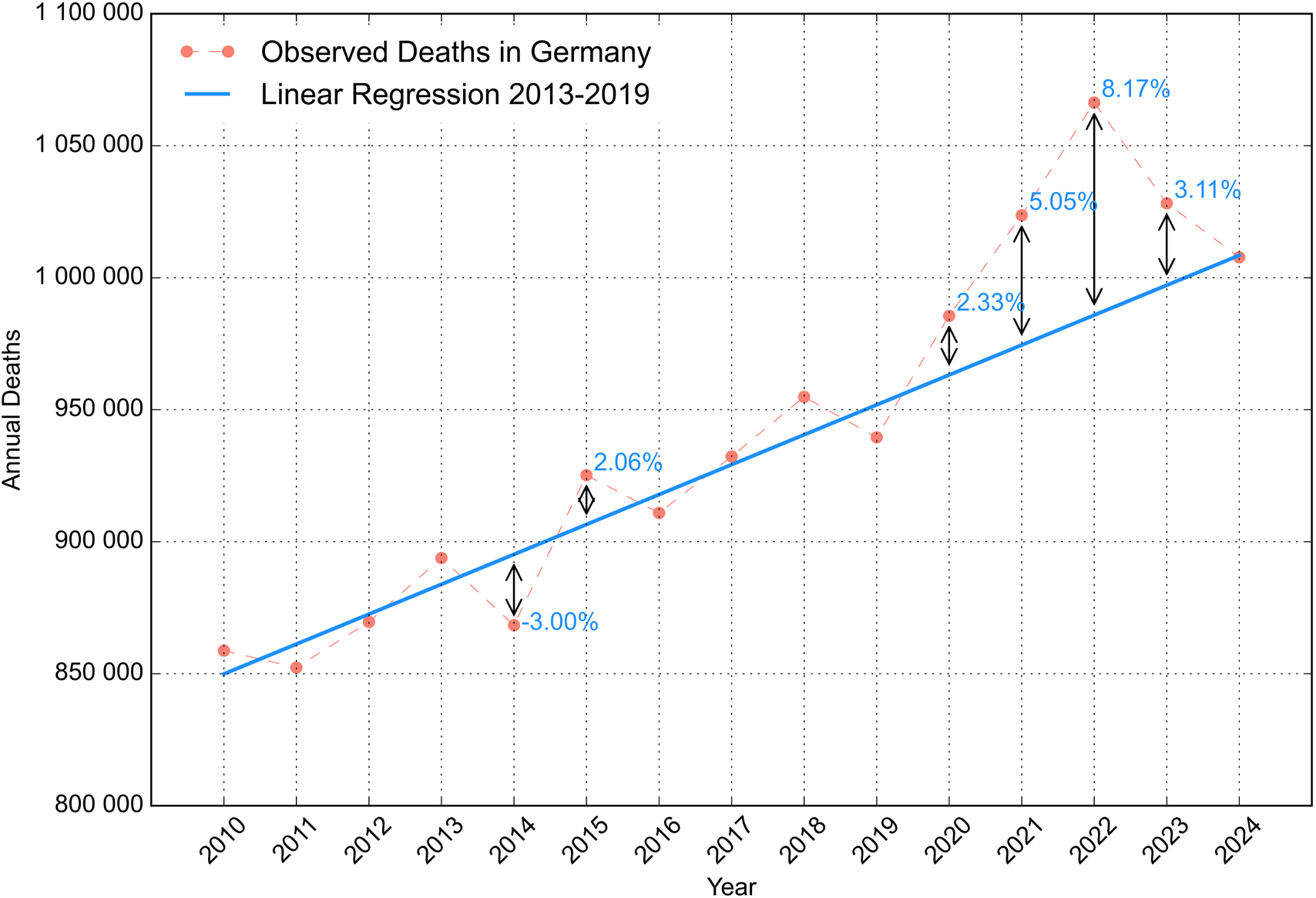
Observed annual deaths in Germany from 2010 to 2024 and linear baseline fitted to the reference period 2013–2019. Relative deviations from the fitted baseline are indicated for the largest negative and positive deviations during the reference period and for the pandemic years 2020–2023.

From 2020 onwards, observed deaths exceeded the extrapolated baseline in all four years investigated. Relative excess mortality amounted to 2.33% in 2020 and increased to 5.05% in 2021 and 8.17% in 2022. In 2023, it declined to 3.11%. The comparison with the deviations within the reference period serve as a simple indicator for the severity of excess mortality. The deviation in 2020 was therefore close to the largest positive deviation observed during the 2013–2019 reference period, whereas the deviations in 2021 and 2022 were substantially larger. In 2023, relative excess mortality had declined markedly compared with 2022 but remained above the deviations observed during the reference period. Mortality in 2024 returned close to the extrapolated pre-pandemic trend.

Absolute and relative values of calculated excess deaths are shown in Table 1 in the first row.

**Table 1.** Absolute and relative excess mortality derived from the 2013–2019 linear baseline, shown for the total population and broad age groups. The last column gives the minimum and maximum relative deviations from the fitted baseline observed during the 2013–2019 reference period. Red values indicate P-scores exceeding the corresponding maximum deviation observed during the reference period.

| **Age Group** | **2020** |  | **2021** |  | **2022** |  | **2023** |  | Min./Max. previous years |
|---|---|---|---|---|---|---|---|---|---|
|  | *absolute* | *relative* | *absolute* | *relative* | *absolute* | *relative* | *absolute* | *relative* |  |
| **Total** | 22 408 | 2.33% | 49 195 | 5.05% | 80 521 | 8.17% | 31 057 | 3.11% | -3.00% / +2.06% |
| **80+** | 24 553 | 4.43% | 35 583 | 6.28% | 63 463 | 10.94% | 23 154 | 3.91% | -3.77% / +2.22% |
| **60-79** | -1716 | -0.53% | 10 219 | 3.18% | 15 277 | 4.76% | 9 378 | 2.93% | -2.14% / +1.97% |
| **40-59** | 168 | 0.23% | 3 763 | 5.25% | 1 717 | 2.43% | -1 029 | -1.48% | -1.81% / +1.43% |
| **20-39** | -71 | -0.73% | 41 | 0.42% | 388 | 3.95% | -51 | -0.52% | -2.92% / +3.61% |
| **0-19** | -522 | -11.29% | -409 | -8.77% | -322 | -6.84% | -389 | -8.19% | -4.05% / +7.65% |

The death-count-based model therefore provides the reference estimates of relative excess mortality against which the ASMR-based estimates are compared in the following analysis.

The corresponding ASMR-based analysis is shown in Figure 2. Age-standardised mortality rates were calculated using 5-year age bands, with 85+ as the open-ended age group and the German population in 2020 (DE2020) as the standard population. In contrast to the increasing trend in annual death counts, ASMR decreased during the 2013–2019 reference period. Relative ASMR-based excess mortality amounted to 2.24% in 2020, 4.74% in 2021, 7.98% in 2022, and 3.41% in 2023. These values closely correspond to the death-count-based estimates of 2.33%, 5.05%, 8.17%, and 3.11%, respectively. Both approaches therefore show the same temporal pattern, with increasing excess mortality from 2020 to a maximum in 2022, followed by a marked decline in 2023.

**Figure 2.**
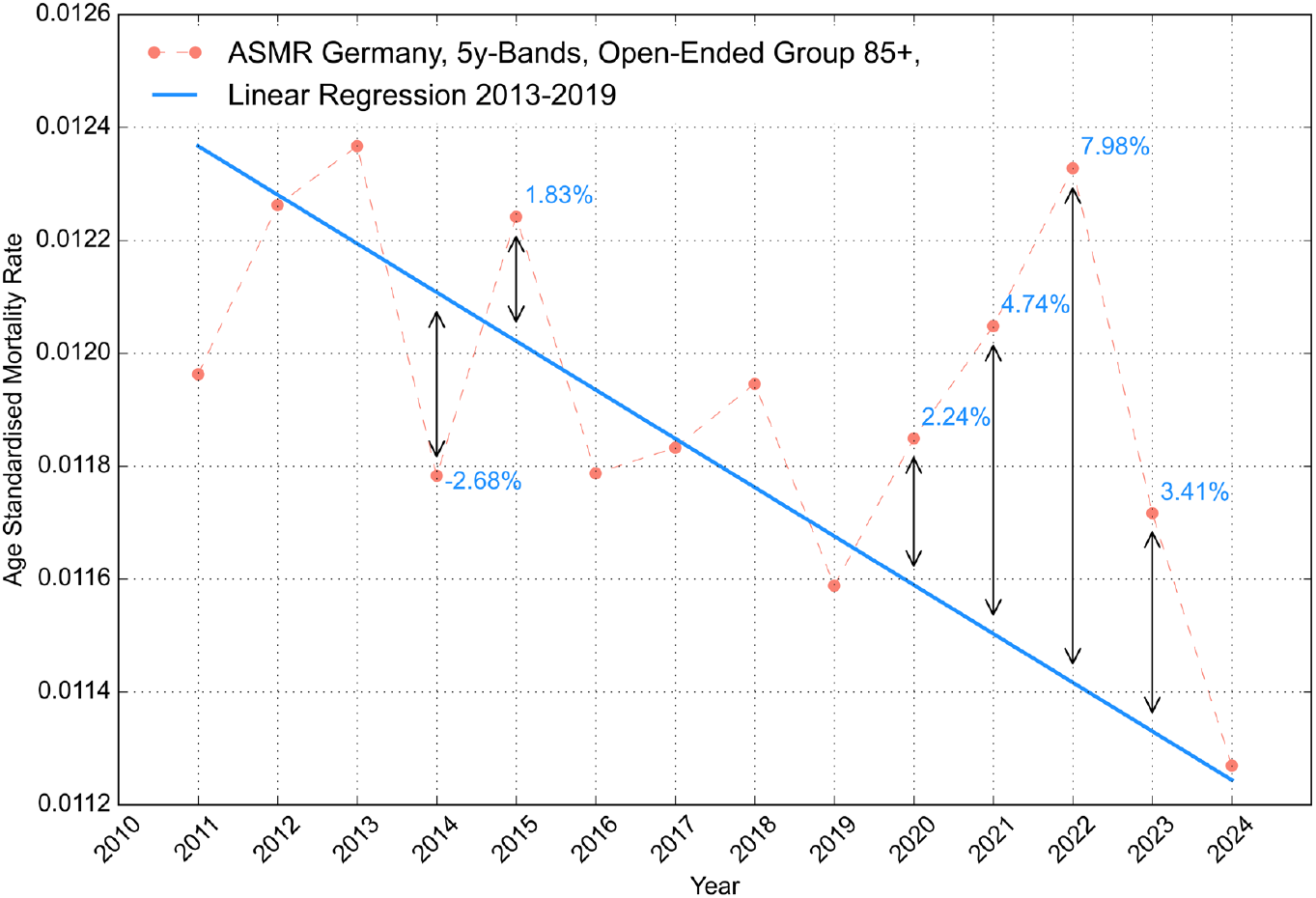
Age-standardised mortality rate (ASMR) in Germany from 2010 to 2024, expressed as annual deaths per person (e.g., 0.012 corresponds to 12 deaths per 1,000 population per year). The German population in 2020 (DE2020) was used as the standard population, with 5-year age bands and 85+ as the open-ended age group. The linear baseline was fitted to 2013– 2019. Relative deviations from the fitted baseline are indicated for the largest negative and positive deviations during the reference period and for 2020–2023.

Within the 2013–2019 reference period, deviations of the observed ASMR from the fitted trend ranged from −2.68% to +1.83%. As for the death-count-based analysis, the deviations observed during 2021 and 2022 were substantially larger than those observed during the reference period. In 2024, the ASMR returned close to the extrapolated pre-pandemic trend.

The close agreement between the two approaches is consistent with the analytical relationship derived in Section 2.5. The remaining differences are examined in the following sections with respect to the choice of standard population, the definition of the open-ended age group, and the baseline model.

#### 3.1.2 Decomposition of Death-Count-Based Excess Mortality by Age

To facilitate the interpretation of the effects discussed in the following sections, death-count-based excess mortality was decomposed by age. For clarity, broader age bands of 20 years were used for this analysis, with 80+ as the open-ended age group. The resulting age-specific P-scores are shown in Table 1, together with the minimum and maximum deviations observed during the 2013–2019 reference period. P-scores exceeding the maximum deviation observed within the respective age group during the reference period are highlighted in red, indicating mortality above the range of deviations observed from the fitted baseline during 2013–2019.

The age distribution of excess mortality differed markedly between the investigated years. In 2020, a P-score exceeding the corresponding maximum deviation during the reference period was observed only in the 80+ age group. In 2021, this pattern extended to the 60–79 and 40–59 age groups. In 2022, P-scores above the respective reference-period maxima were observed across all age groups from 20 years onwards, whereas in 2023 this was restricted to the two oldest age groups. Thus, excess mortality was strongly concentrated in the oldest age group in 2020, while in 2022 it was distributed much more broadly across age groups.

### 3.2 Effect of the Standard Population

To investigate the effect of the standard population, ASMR-based excess mortality was calculated using three different standard populations while keeping the age-group definition and all other parameters unchanged. Five-year age bands were used, with 85+ as the open-ended age group. The German population in 2020 (DE2020), the European Standard Population 2013 (ESP2013), and the WHO standard population (WHO) were used. The resulting P-scores are shown in Table 2.

**Table 2.** ASMR-based relative excess mortality (P-scores) for 2020–2023 using the German population in 2020 (DE2020), the European Standard Population 2013 (ESP2013), and the WHO standard population. All calculations use the same age-group definition and the 2013–2019 reference period. The last column gives the minimum and maximum relative deviations from the corresponding fitted ASMR baseline observed during the reference period.

| **Standard Population** | **2020** | **2021** | **2022** | **2023** | **min./max.** |
|---|---|---|---|---|---|
| **DE2020** | 2.24% | 4.74% | 7.98% | 3.41% | -2.68%/1.83% |
| **ESP2013** | 2.26% | 4.82% | 7.95% | 3.27% | -2.69%/1.73% |
| **WHO** | 1.81% | 5.39% | 7.93% | 3.86% | -2.54%/1.64% |

The choice of standard population had the strongest effect in 2020. The P-score changed from 2.24% using DE2020 to 2.26% using ESP2013 and decreased to 1.82% using WHO. This is consistent with the age distribution of excess mortality shown in Table 1: in 2020, excess mortality was concentrated in the oldest age group. Standard populations assigning relatively less weight to the oldest age groups therefore result in a lower overall estimate of relative excess mortality.

In 2022, when excess mortality was distributed more broadly across age groups, the dependence on the standard population was considerably smaller. The corresponding P-scores ranged only from 7.98% to 7.95% using ESP13 and 7.93% using WHO. For 2021 and 2023, the differences were also relatively small, although they were not monotonic with the age structure of the standard population. Despite these quantitative differences, all three standard populations yielded the same qualitative result: for each year from 2020 to 2023, the P-score exceeded the maximum deviation observed during the corresponding 2013–2019 reference period. Thus, the effect of the standard population depends not only on the age structure of the standard population itself, but also on the age-specific distribution of excess mortality in the year under consideration.

### 3.3 Effect of Age Bands and the Open-Ended Age Group

In addition to the choice of standard population, the definition of the age groups affects ASMR-based excess mortality estimates. To investigate this effect independently of the standard population, all calculations in this section use DE2020 as the standard population. Table 3 compares calculations using 5-year age bands with 85+ or 80+ as the open-ended age group, as well as 10-year age bands with 80+ as the open-ended age group.

**Table 3.** ASMR-based relative excess mortality (P-scores) obtained with different age-group definitions using DE2020 as the standard population. Five-year age bands with open-ended groups of 85+ and 80+ are compared to isolate the effect of the upper age-group definition; 5- and 10-year age bands with the same open-ended group of 80+ are compared to assess the effect of age-band width. All calculations use the 2013–2019 reference period. Minimum and maximum values refer to deviations from the corresponding fitted baseline observed during the reference period.

| Age Bands / lower boundary of open-ended age group | 2020 | 2021 | 2022 | 2023 | min./max. |
|---|---|---|---|---|---|
| **5y / 85+** | 2.24% | 4.74% | 7.98% | 3.41% | -2.68%/1.83% |
| **5y / 80+** | 1.82% | 4.83% | 8.97% | 5.78% | -2.56%/2.34% |
| **10y / 80+** | 1.09% | 3.39% | 6.78% | 3.25% | -2.62%/2.34% |

Changing the open-ended age group from 85+ to 80+, while retaining 5-year age bands for all younger ages, resulted in substantial differences in the estimated P-scores. In 2020, the P-score decreased from 2.24% to 1.82%, whereas in 2022 it increased from 7.98% to 8.97% and in 2023 from 3.41% to 5.78%.

The effect is particularly relevant for 2020: using 85+ as the open-ended age group, the P-score of 2.24% exceeded the maximum deviation of 1.83% observed during the corresponding reference period. With 80+ as the open-ended age group, however, the P-score of 1.82% remained below the corresponding reference-period maximum of 2.34%. Thus, the definition of the open-ended age group changes the qualitative assessment of mortality in 2020 relative to the deviations observed during the reference period. Thus, combining the 80–84 and 85+ age groups does not result in a systematic upward or downward shift of the estimated excess mortality, but changing age bands from 5 years to 10 years while maintaining the open ended group at 80 years and more moves P-scores down for all years under investigation.

The origin of this effect is illustrated in Figure 3. The figure compares the contribution of the population aged 80 years and older to the ASMR when the 80–84 and 85+ age groups are standardised separately (stratified calculation) with the contribution obtained when 80+ is treated as a single open-ended age group (aggregated calculation). Both calculations cover exactly the same population and use DE2020 as the standard population.

**Figure 3.**
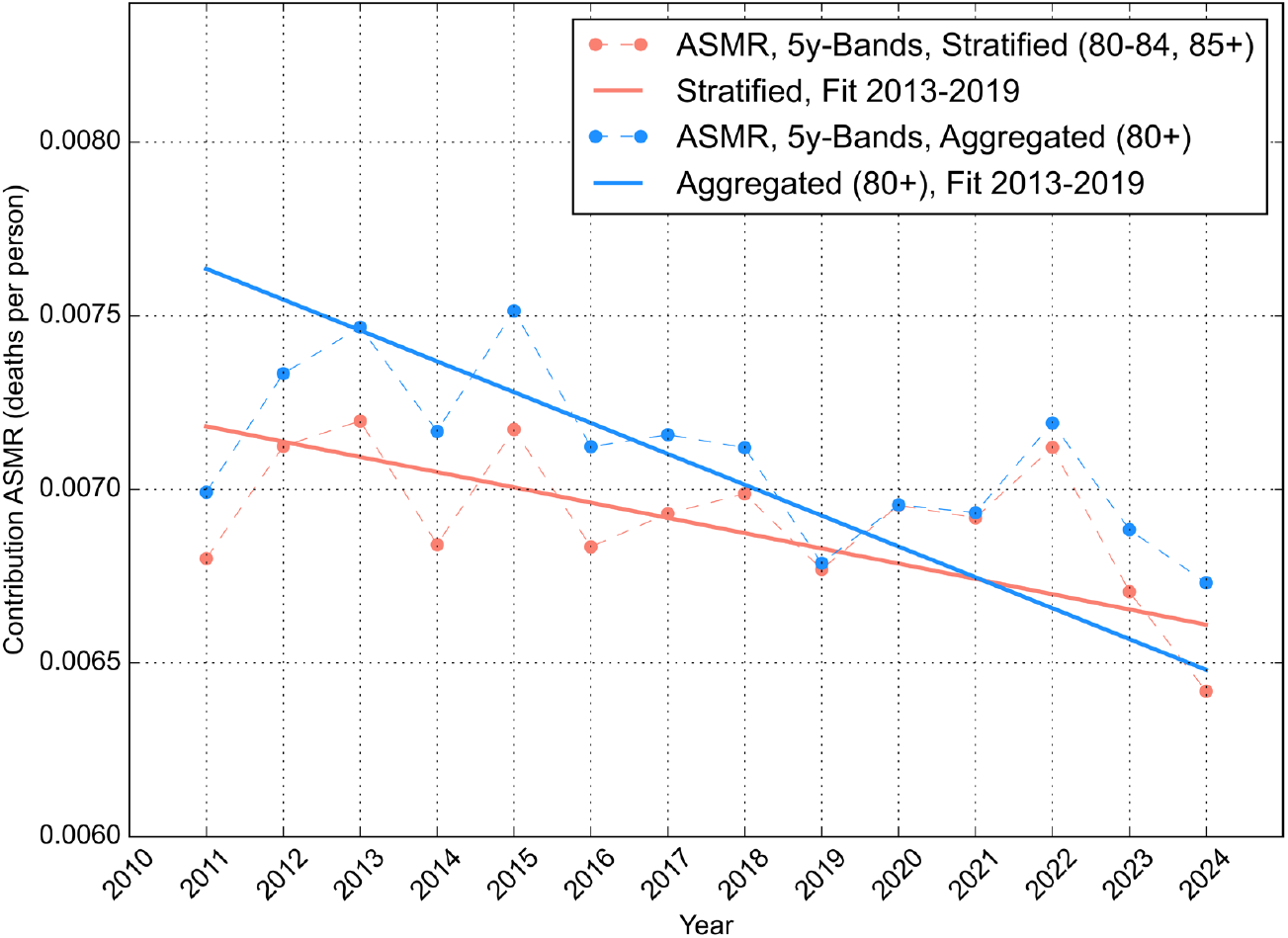
Contribution of the population aged 80 years and older to the age-standardised mortality rate using DE2020 as the standard population. In the stratified calculation, the 80–84 and 85+ age groups are standardised separately and their contributions are summed; in the aggregated calculation, 80+ is treated as a single open-ended age group. Linear fits to 2013– 2019 demonstrate that aggregation changes the pre-pandemic trend despite both calculations covering the same population aged 80 years and older.

Nevertheless, the two approaches produce different ASMR contributions during the 2013–2019 reference period and, consequently, different linear trends. In the aggregated calculation, changes in the relative proportions of individuals aged 80–84 and 85+ remain within the mortality rate of the 80+ group. In the stratified calculation, these changes in age composition are controlled by standardising the two age groups separately. Aggregation therefore changes the pre-pandemic ASMR trend and, consequently, the extrapolated expected mortality and the resulting P-score.

A further effect is observed when the age-band width is increased from 5 to 10 years while retaining 80+ as the open-ended age group. The resulting P-scores are lower in all four investigated years (Table 3). Thus, ASMR-based excess mortality is affected not only by the choice of standard population, but also by the age-group definition used for standardisation.

### 3.4 Analytical calculation of the Effect of Aggregation in the Open-Ended Group

The observed aggregation effect in the former section can be explained analytically: Consider two age groups (let the first be the younger one e.g. 80-84 and the second one the older one e.g. 85+) with death counts *d*_*1*_ and *d*_*2*_, population sizes *p*_*1*_ and *p*_*2*_ and corresponding weights *s*_*1*_ and *s*_*2*_ in the standard population.

The aggregated and the stratified contribution to the ASMR are:

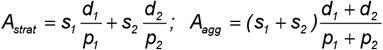

Their difference can be written and rearranged:

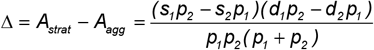

As all of the values of *d*_*i*_, *p*_*i*_ and *s*_*i*_ are positive, the sign of Δ is determined by the expression:

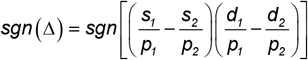

And there are interesting consequences for this application here.

First, if the weights of the standard population are equal to the weights of the population under investigation (like here for 2020) the expression becomes zero and *A*_*agg*_ *( 2020 )* = *A*_*strat*_ *( 2020 )* . This is visible for 2020 in Figure 3.

For the years before 2020, the population was younger, so the proportion of the younger group was higher than the proportion of the older one, so:

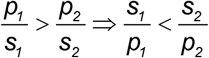, making the sign of the first factor negative. For the second factor, the sign of the expression is always negative, as mortality increases with age. This makes *Δ* become positive and *A*_*strat*_ always bigger than *A*_*agg*_ for the years before 2020.

For the population development considered here, this results in a steeper temporal increase of the aggregated contribution than of the stratified contribution: *m*_*agg*_ *> m*_*strat*_.

Thus, age-group aggregation can affect not only the level of an ASMR but also its temporal trend. When this trend is extrapolated to estimate expected mortality, the choice of the open-ended age group can therefore alter the expected ASMR and, consequently, the estimated P-score.

### 3.5 Effects of Baseline Selection

Finally, the effect of baseline selection was investigated by repeating the death-count- and ASMR-based calculations using six different pre-pandemic reference periods, ranging from 2011–2019 to 2016–2019. For the ASMR calculations, DE2020 was used as the standard population with 5-year age bands and 85+ as the open-ended age group. Figure 4 compares the resulting death-count- and ASMR-based P-scores for each reference period and for the years 2020–2023. The dashed identity line indicates equal P-scores obtained from both approaches.

**Figure 4.**
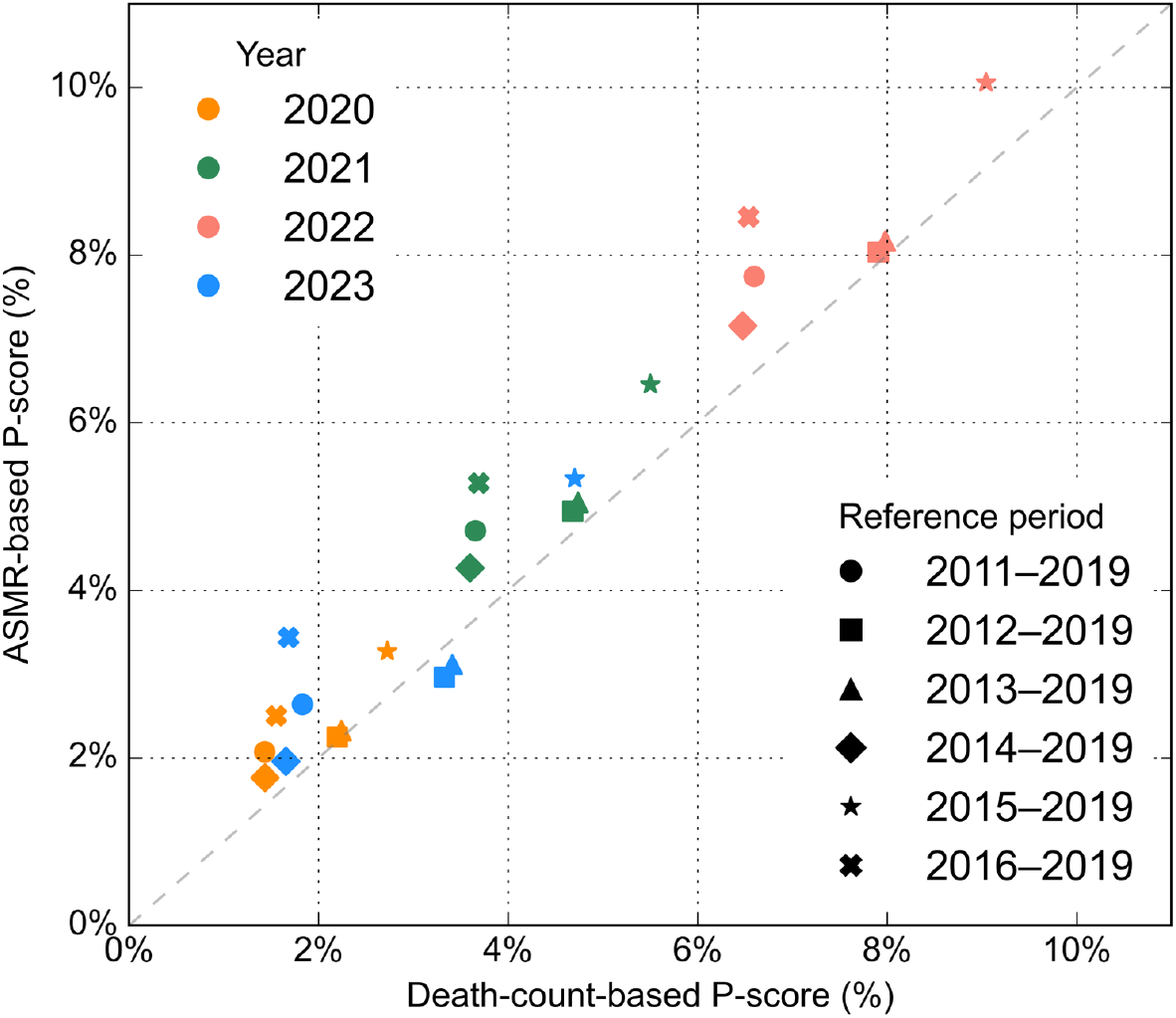
Relationship between death-count-based and ASMR-based P-scores for different pre-pandemic reference periods. Colours indicate the years 2020–2023 and symbols indicate the reference periods used for linear baseline estimation. ASMRs were calculated using DE2020 as the standard population, 5-year age bands, and 85+ as the open-ended age group. The dashed line indicates equality between death-count- and ASMR-based P-scores.

Baseline selection had a substantial effect on the estimated magnitude of excess mortality. Nevertheless, both approaches responded systematically to changes in the reference period: baseline specifications producing higher death-count-based P-scores also produced higher ASMR-based P-scores. The 2012-2019 and 2013–2019 reference periods, which yield estimates approximately in the middle of the range obtained from the investigated baseline specifications, also produced the closest agreement between both measures, with the corresponding P-scores located close to the identity line. In contrast, the shorter 2015–2019 reference period produced markedly higher P-scores with both approaches, the reference period 2014-2019 produces lowest P-scores for both quantities.

For the other investigated reference periods, ASMR-based P-scores were consistently higher than the corresponding death-count-based estimates. Thus, baseline selection affects not only the magnitude of estimated excess mortality but also the quantitative agreement between the two approaches.

Nevertheless, the systematic co-variation of both measures demonstrates that the choice of the pre-pandemic baseline is an important determinant of excess mortality estimates irrespective of whether they are derived from death counts or ASMR.

## 4 Summary and Discussion

### 4.1 Relationship between Death-Count- and ASMR-Based Excess Mortality

The analytical and empirical results demonstrate a close relationship between excess mortality derived from death counts and from age-standardised mortality rates. Death-count-based relative excess mortality can be expressed as a weighted combination of age-specific excess mortality, with the expected numbers of deaths defining the weights. ASMR-based excess mortality modifies these weights according to the relationship between the age distribution of the standard population and that of the population under study. If both distributions are identical, both approaches yield the same relative excess mortality.

This relationship is reflected in the German data. Although annual death counts increased during the pre-pandemic period while ASMR decreased, both approaches produced similar P-scores for 2020– 2023. Death counts therefore provide a comparatively simple and transparent measure of relative excess mortality when a single population is analysed over time. The analytical relationship clarifies under which conditions both approaches provide similar results and why they may differ.

### 4.2 Effects of the Standard Population and Age-Group Definition

The choice of standard population affects ASMR-based excess mortality through the weighting of age-specific mortality. The magnitude of this effect depends on the age distribution of excess mortality itself. This explains the larger sensitivity observed in 2020, when excess mortality was strongly concentrated in the oldest age group, compared with 2022, when elevated mortality was distributed much more broadly across adult age groups. The changing age pattern of excess mortality during the pandemic has also been demonstrated in the underlying age-stratified analysis.

The definition of the age groups introduces an additional and less obvious effect. Aggregating 80–84 and 85+ into a single 80+ group leaves changes in age composition within this group unaccounted for by standardisation. Because mortality differs substantially between 80–84 and 85+, such compositional changes affect the mortality rate of the aggregated group. In contrast, when both groups are standardised separately, their relative weights are fixed by the standard population.

This distinction is particularly relevant for trend-based excess mortality estimates. With DE2020 as the standard population, the aggregated and stratified calculations coincide when the age composition corresponds to that of 2020. In earlier years, however, the population aged 80+ was relatively younger. Consequently, aggregation changes the pre-pandemic ASMR trend and therefore the extrapolated expected mortality. The resulting difference in the P-score is thus not primarily caused by the observed mortality in the target year, but by a different baseline.

This finding highlights an interaction between standard population and age-group definition. A standard population can control changes in age structure only at the resolution provided by the selected age groups. For trend-based excess mortality estimation, the standard population should therefore not be regarded merely as a scaling convention; together with the age-group definition, it determines which demographic changes are removed by standardisation and which remain in the mortality trend.

### 4.3 Impact of Baseline Selection and Methodological Implications

Baseline selection had a substantial influence on the magnitude of estimated excess mortality. However, death-count- and ASMR-based P-scores responded in a closely corresponding manner when the reference period was varied. The 2013–2019 baseline produced estimates approximately in the middle of the range obtained from the alternative reference periods. This agrees with the previous sensitivity analysis, in which the 2013–2019 death-count trend yielded the median projected value and a slope close to the average of the investigated trends.

The 2015–2019 reference period provides a useful counterexample, producing markedly higher excess mortality estimates with both approaches.

Taken together, the results identify three closely related methodological choices affecting estimates of relative excess mortality: the baseline used to estimate expected mortality, the standard population used for age standardisation, and the age-group definition. Death-count-based analysis avoids the latter two choices and therefore provides a useful transparent reference measure for analyses within a single population. ASMR additionally controls for demographic change but introduces assumptions concerning standard population and age-group resolution that need to be reported and considered when interpreting trend-based excess mortality estimates.

The empirical analysis is limited to Germany and to the mortality patterns observed during 2020–2023. The magnitude of the effects identified here may differ in populations with other age structures or mortality patterns. Furthermore, the empirical calculations use linear baseline models, although the analytical relationships between death-count- and ASMR-based excess mortality are not restricted to the German pandemic period. Population estimates used as denominators for age-specific mortality rates are themselves subject to demographic estimation and census revisions, providing an additional source of uncertainty in age-standardised analyses.

## 5 Conclusions

Death-count- and ASMR-based estimates of relative excess mortality are analytically closely related and can yield equivalent results under defined conditions. The German data demonstrate that differences between the two approaches depend systematically on the standard population, the age-group definition, and the baseline used to estimate expected mortality.

In particular, aggregation of the oldest age groups can alter the pre-pandemic ASMR trend by retaining changes in age composition within the open-ended age group. Consequently, age-group definition can affect not only the level of an age-standardised mortality rate but also the resulting estimate of excess mortality.

For analyses within a single population over time, death counts therefore provide a simple and transparent reference measure. More generally, excess mortality estimates should be interpreted in the context of the baseline, standard population, and age-group definition from which they were derived.

## Data Availability

https://github.com/MartinSauter/Death_counts_vs_ASMR_Germany

## 6 Declaration of ethics, funding and personal interests

This publication uses only publicly available data, so there was no need to consult an ethic commission. I declare that I did not receive any funding for this work and that I have no concurring personal interests. Clinical trial number: not applicable

## 7 Data and Code Availability

Data and Python code for this study are available under https://github.com/MartinSauter/Death_counts_vs_ASMR_Germany

### 8 Abbreviations

CMR: Crude Mortality Rate
CDC: Crude Death Counts
ASMR: Age-standardised Mortality Rate
Destatis: Deutsches Statistisches Bundesamt
HMD: Human Mortality Database
WMD: World Mortality Dataset
NHS: National Health Service

## Notes

### Competing Interest Statement

The authors have declared no competing interest.

### Author Declarations

This publication uses only publicly available data, so there was no need to consult an ethic commission.

